# Keeping the ‘C’ in CBPR: Exploring Community Researchers’ Experiences with Human Subjects Protection Training Requirements

**DOI:** 10.1101/2023.03.23.23287595

**Authors:** Naomi Cruz, Christiana Adams, Constance Akhimien, Fauziyya Allibay Abdulkadir, Cherriece Battle, Maria Oluwayemi, Olanike Salimon, Teri Lassiter, Leslie Kantor

## Abstract

Community-engaged research is increasingly recognized for its potential to advance health equity. The ability to conduct such research in the United States is predicated on the completion of human subjects protection courses; however, prior studies suggest that many of these required courses may not adequately accommodate the varied skillsets and backgrounds of community members involved with carrying out research. The present study explores community researchers’ (CRs’) experiences with a widely used human subjects protection course frequently mandated by universities. Six CRs involved in conducting a community-based participatory research (CBPR) project on Black women’s pregnancy-related experiences were interviewed about their completion of a required human subjects protection course. Across multiple interviews, CRs noted challenges with the training length, competing external demands, module readability, content relevancy, end-of-module quizzes, and technology requirements. Despite such obstacles, CRs still valued the opportunity to learn and felt more knowledgeable and capable post-course completion. Recommendations for course improvement were explored. University requirements for human subjects protection trainings may place an undue burden on community members preparing to conduct research, impede academic-community partnerships, and discourage the initiation and continuation of community-engaged studies. Course alternatives that are tailored to CRs are needed.

## Introduction

Community-based participatory research (CBPR) centers on the equitable and active involvement of community members throughout all stages of the research process.^1,2^ Grounded in the principles of social justice, CBPR is designed to empower people of various backgrounds, skillsets, and research experiences to draw upon their unique understandings of local contexts and convert their desires for change into action.^3–5^ The promises of CBPR lie largely in its potential to elevate the voices of community members who have been historically marginalized by situating such individuals as equal partners to academic researchers in the planning, execution, analysis, and dissemination phases of qualitative studies. CBPR’s mission to conduct research *with* communities rather than *on* communities is captured in the nine core principles upon which it is based.^6^ These include, for example, fostering co-learning among all research partners, engaging in iterative processes, and disseminating subsequent findings and knowledge to communities in an appropriate and accessible manner with the ultimate aim of effecting meaningful and long-lasting change.^6^

Prior studies support CBPR as a lever for improving health equity.^7–9^ For instance, populations that have been historically mistreated in research may be more willing to engage with fellow community members than with researchers from academic institutions.^5,10^ Equitably partnering with community members in crafting and implementing research can be valuable both in nurturing trust and developing public health interventions which are evidence-based, feasible, relevant, and effective.^11,12^ Moreover, CBPR can strengthen community-academic partnerships; bolster community members’ ability to explore and advocate for solutions to relevant health problems; influence health policy; and increase researchers’ access to and understanding of local insights, attitudes, and information.^5,13,14^

For community members and academics alike, the ability to conduct CBPR in the United States is predicated on the completion of human subjects protection courses intended to safeguard the “rights, welfare and safety” of research participants.^15^ The most extensively utilized human subjects protection courses among U.S. academic institutions are those provided by the Collaborative Institutional Training Initiative (CITI) Program, an entity which “identifies education and training needs in the communities [it serves] and provides high quality, peer-reviewed, web-based educational materials to meet those needs.”^16,17^

The availability of standardized trainings on human subjects protection is intended to equip all who are involved in the research process with the tools they need to safeguard participants’ well-being. However, previous studies suggest that such trainings may fail to adequately accommodate the varied skillsets, educational backgrounds, and scientific experiences of community members involved with conducting human subjects research.^18–20^

The present investigation constitutes a sub-study within a broader CBPR project on Black women’s pregnancy-related experiences in a city with high rates of maternal mortality and severe maternal morbidity. As part of this project, the university-affiliated authors partnered with six community members who joined the study team (referred to as “community researchers,” or “CRs,” from here forward). In this paper, the full research team examines CRs’ experiences with completing a required CITI Program human subjects course and their recommendations for change.

## Methods

CRs were identified in conjunction with a local nonprofit, the lead grantee on the overarching maternal health project. Outreach efforts to advertise this opportunity included disseminating flyers to the nonprofit’s clinical and community partners, posting on social media, and messaging current clients. As community members found out about the opportunity, they also circulated information within their own social networks. All community members who joined the study team were paid for their work on the research project.

Prior to beginning the mandated human subjects course, CRs participated in a tailored, four-part training series developed and delivered by university researchers involved in the project. These preparatory sessions included both didactic presentations and interactive exercises on the principles of CBPR; developing open-ended questions; balancing self-disclosure with research objectives; and research ethics along with the CITI Program’s Social / Behavioral / Epidemiologic Investigators Course, the human subjects course utilized by the university. The final session was formulated to help CRs prepare for and gain familiarity with the required human subjects course. This session consisted of a two-hour training which included how to engage with the course’s online interface; test-taking tips; practice quiz questions; and other relevant information, skills, and strategies related to the mandated course. Working with the university research team, CRs also generated and refined an interview guide for the larger maternal health study. The four tailored training sessions were conducted synchronously via Zoom over the span of approximately one month. Following the final session, CRs were instructed to complete the human subjects course in preparation for the CBPR project.

All CRs were invited to and agreed to participate in a sub-study about their experience with the CITI Program. The broader CBPR study from which this sub-study is derived was approved by the University’s IRB (Pro2019002981). The present sub-study was determined to be non-human subjects research. Semi-structured interviews with CRs ranged from 15 to 30 minutes and were based on a question guide. The guide covered topics including overall opinions about the CITI Program; course length, readability, content, and quizzes; applying CITI teachings to CR fieldwork; technology requirements; and ideas for course improvement.

CRs completed the CITI course in November 2021. Individual interviews were conducted via Zoom between December 2021 and March 2022. Permission to record the interviews was granted by each CR at the time of the interview. A general inductive approach was employed in order to identify themes across the six interviews.^21^ Each Zoom-generated transcript was coded. The resulting codes were then grouped into themes and analyzed.

## Results

Several key themes and recommendations based on CRs’ experiences with the required human subject training emerged across the six interviews. Additional CR recommendations for a tailored and improved human subjects protection course are included in Table 1.

**Table 1.**
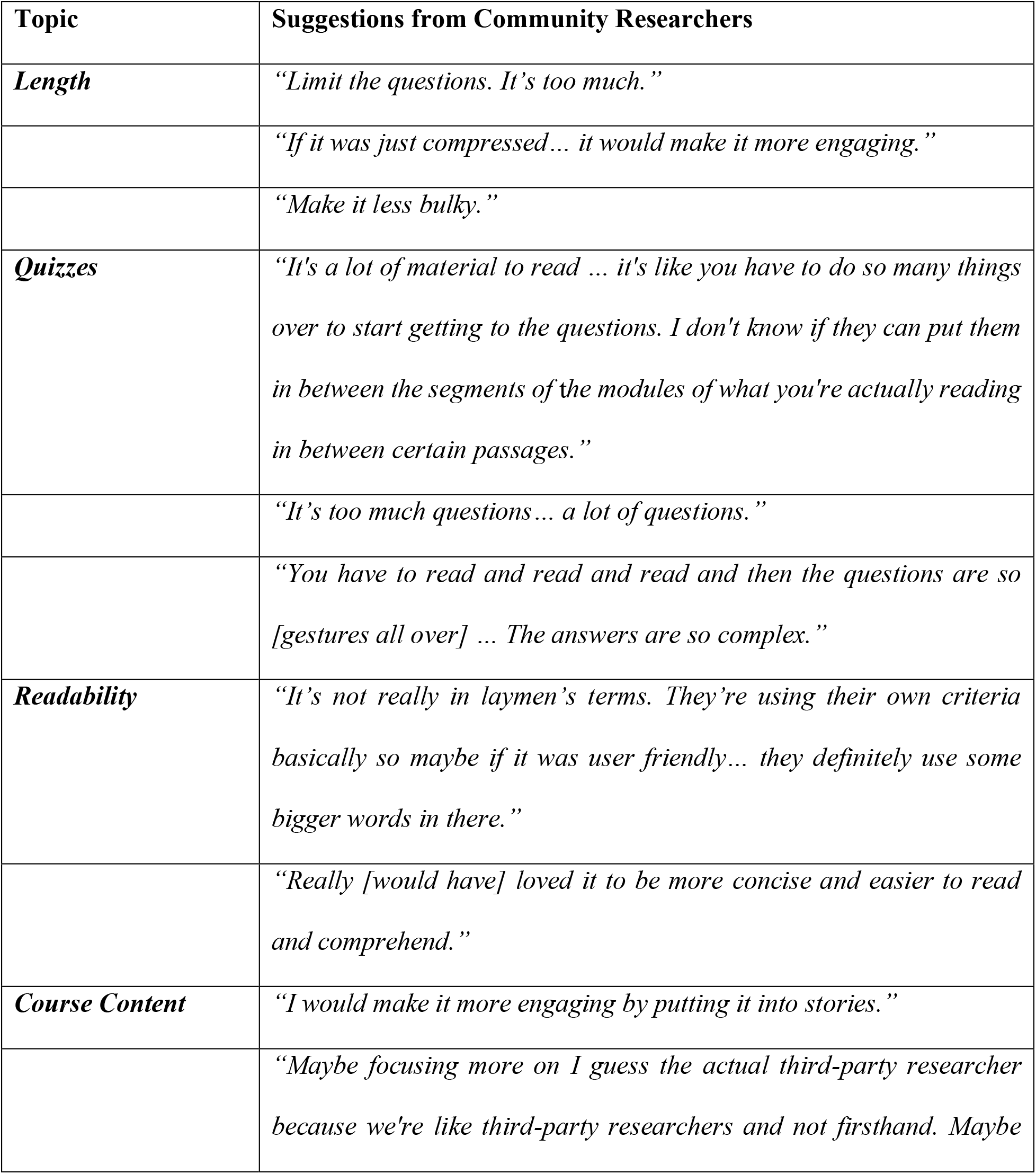
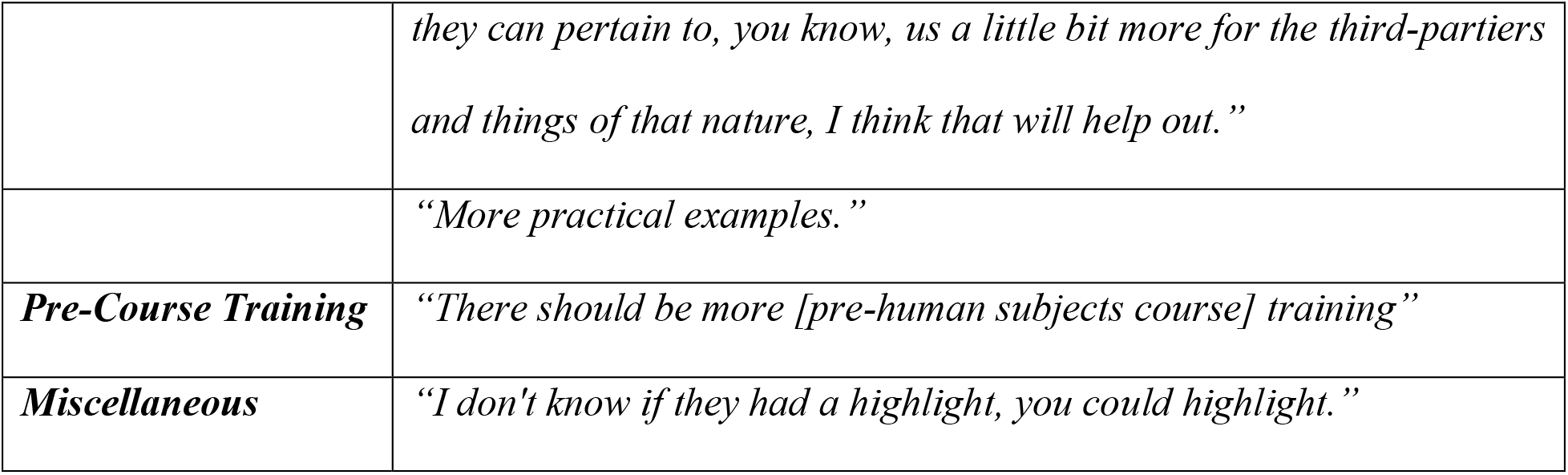
Community Researcher Recommendations on How to Improve the Social / Behavioral / Epidemiologic Investigators Course and Better Tailor the Curriculum to Community Researchers.

| **Topic** | **Suggestions from Community Researchers** |
| --- | --- |
| ***Length*** | *“Limit the questions. It’s too much.”* |
|  | *“If it was just compressed... it would make it more engaging.”* |
|  | *“Make it less bulky.”* |
| ***Quizzes*** | *“It’s a lot of material to read ... it’s like you have to do so many things over to start getting to the questions. I don’t know if they can put them in between the segments of the modules of what you’re actually reading in between certain passages.”* |
|  | *“It’s too much questions... a lot of questions.”* |
|  | *“You have to read and read and read and then the questions are so [gestures all over] ... The answers are so complex.”* |
| ***Readability*** | *“It’s not really in laymen’s terms. They’re using their own criteria basically so maybe if it was user friendly... they definitely use some bigger words in there.”* |
|  | *“Really [would have] loved it to be more concise and easier to read and comprehend.”* |
| ***Course Content*** | *“I would make it more engaging by putting it into stories.”* |
|  | *“Maybe focusing more on I guess the actual third-party researcher because we’re like third-party researchers and not firsthand. Maybe* |
|  | *they can pertain to, you know, us a little bit more for the third-partiers and things of that nature, I think that will help out.”* |
|  | *“More practical examples.”* |
| ***Pre-Course Training*** | *“There should be more [pre-human subjects course] training”* |
| ***Miscellaneous*** | *“I don't know if they had a highlight, you could highlight.”* |

### Training length

The most widely discussed challenge across all interviews was the length of time required to complete the 15-module course.

Each of the six CRs remarked on the “bulkiness” of the training. The majority emphasized that, even when they perceived the material to be interesting or informative, the large number and lengthy nature of the modules led to reduced levels of engagement and concentration as they progressed through the program. One CR noted that the amount of time required to finish this course made it feel as though she had taken on a “full-time job” or “PhD program.” The extensive task of completing the training was often viewed as tedious, as is suggested by one CR’s comment: “let me see how many pages are left. Oh, it’s not ending.”

### Balancing the required course with external demands

Another theme involved the need to balance completing the human subjects protection course with external demands. Although the tailored training sessions developed by the university researchers allotted some time for CRs to work on the course, nearly all CRs needed multiple hours of additional time to finish.

CRs often worked on the university-required training while managing childcare and work responsibilities. As one CR explained: “Some mothers don’t even have that luxury of time to sit down for so much like that because they have a lot of things to take care of… Like me, it took me a while because, you know, I’m too busy. I’m the only one, my husband isn’t here, so I just feel it’s [too] much for me you know?… It’s too much for me because I have to take care of a lot of things.”

Other CRs felt that they were able to balance training demands and external obligations without issue. These CRs generally cited having a clear deadline by which they needed to complete the training, employing time management skills, and adopting other proactive measures as helpful in meshing their research commitments with their non-research-related responsibilities.

### Readability

There were mixed perceptions regarding how challenging it was to comprehend the modules and quizzes. Some CRs reported no issues, with one individual referring to it as “basic comprehension,” while others reflected on jargon that hindered their ability to understand the material. One CR stressed that the training would have been improved if CITI had opted to use “simple text, straight to the point of what they’re trying to bring out,” adding later that “they make it tactical, like some kind of puzzle, but we don’t need puzzles.”

### Quizzes

CRs expressed mixed emotions about the quizzes, ranging from frustrated and stressed to unphased and appreciative of select quiz functions.

A repeated critique of the quizzes was that no matter how thoroughly one read through or listened to the module, the quiz questions and answers were so complex that even the most well-prepared test-taker could read it and still not “get it.” This perceived disconnect between the effort placed into learning the material and subsequent quiz scores caused some CRs to feel frustrated and as though the CITI course was not intended for or applicable to them. As one CR remarked, “the questions were too much for just a community researcher.”

Nevertheless, CRs appreciated the opportunity to retake the quizzes if the minimum grade threshold was not met. Some CRs had trouble understanding what particular quiz questions were asking and had to retake the quizzes several times prior to receiving a passing score. This ability to retake the quizzes helped relieve some of the pressure CRs felt to pass these assessments on their first attempt.

### Relevance of course content to CBPR

CRs indicated that, while course content was informative, it could also be abstract, outdated, and outside the scope of the study they were preparing to conduct.

One CR commented that the training resembled a history lesson while others stated that the course did not adequately connect human subjects research with effecting positive change in their community. Multiple CRs suggested that insufficient attention was allotted to subject recruitment, fieldwork, and community engagement throughout the course and that more applied, succinct, and true-to-life stories and examples would have been helpful.

Some CRs likewise felt that they were not included in CITI’s target demographic for this course. As one CR voiced: “I didn’t feel like [the modules] pertained to us, specifically for the researcher part…it’s annoying to keep going through the same thing as if we’re in the research, you know, like we’re in the administrative part… It’s almost like we’re the civilians and the course is [for] the head honchos. We’re not really at that point, you know, of doing that.”

It should be noted that CITI does have some modules which specifically address community research; however, these modules are intended to “supplemen[t] the foundational training provided in a basic Human Subjects Research (HSR) course.”^22^ Thus, had these modules been mandated as well, they would have been in addition to the 15 currently-required modules and would have generated an additional burden for CRs.

Despite these critiques, CRs who had begun working with human subjects prior to being interviewed noted that certain modules – in particular, those pertaining to conflicts of interest and confidentiality – increased their knowledge, confidence, and comfort with study participants. CRs who had not yet begun working with human subjects at the time they were interviewed generally cited greater levels of knowledge and confidence as well.

### Technology Requirements

Technology presented a barrier to CRs completing the human subjects training. There were multiple steps required to obtain university log-in credentials, register for the course, enroll in the IRB system, and then link the training completion record to the IRB system. Enlisting the university’s IT department helped resolve select difficulties regarding registration procedures and navigating the CITI interface. CRs repeatedly commented on the value of having the study coordinators and IT to call on when in need of technology-related help, noting how this made the processes “smooth.”

Multiple CRs completed the training on their smartphones and found the ability to do so convenient. These individuals did not report greater difficulties with completing the training than did those who used computers.

### Cultural insensitivity

No CRs felt that the CITI Program was constructed or delivered in a way that was culturally insensitive.

### Valued learning experience

Although CRs reported a number of challenges with the training, there was a shared sentiment among interviewees that “no knowledge is wasted” and that the course helped “broaden [their] horizons.” All CRs conveyed that they “learned a lot,” and a few remarked on the satisfaction that stemmed from completing such a difficult task and having a certificate to prove it. Many expressed that completing the training bolstered their confidence in conducting interviews and in their own capabilities as researchers.

### The impact of preparatory, tailored trainings

University researchers involved in this study developed and delivered a four-session supplemental training which sought to introduce CRs to the fundamentals of research/research ethics, familiarize CRs with the mandated human subjects course, and create opportunities for question-asking. As implied above, the provision of this project- and CR-specific, preparatory training did not fully eliminate the challenges associated with the subsequent human subjects protection course; however, CRs did report that these sessions had a positive impact on their ability to complete the course and on their feelings of self-efficacy as interviewers. Multiple CRs suggested that future research should involve more of such training sessions which preview course material and relate it back to the project for which they are preparing.

In an informal interview conducted after the data collection phase, CRs noted a preference for “training[s] related to what we’re doing” as compared to those containing more generalized curricula. CRs expressed a specific desire to learn about topics directly related to the CBPR project (e.g., maternal health, hospital systems) rather than information with little bearing on the project at hand (e.g., research involving prisoners) which was included in the mandatory training they completed. [Table 1 near here].

## Discussion

Interviews with six community members trained as community researchers reveal that widely required human subjects protection courses may not address CRs’ unique skillsets, backgrounds, and experiences and may instead create impediments to CBPR.

In recalling their experiences, CRs noted problems relating to training length, competing external demands, overall readability, end-of-module quizzes, course content relevancy to CR-specific roles, and technology requirements. Key recommendations for course improvement included reducing the training length, ensuring concordance between modules and quiz questions, increasing material comprehensibility, tailoring the training to reflect CR-specific roles and project-specific aims (e.g., removing unrelated modules, adding a focus on maternal health), and simplifying technology requirements regarding linking course completion records to the university’s IRB system.

Other training attributes revealed by this study may also stand as barriers to community involvement on research teams. For example, while full-time researchers are often accustomed to the academic writing style and complex question structures utilized in many training programs, such language can be intimidating and hard to understand for people who lack research training. As the average reading level for these mandated courses can be far above the national average,^17^ individuals with lower literacy levels may struggle to understand or complete mandatory courses. Accordingly, while the present human subjects training was designed for “all persons involved in research studies involving human subjects,”^23^ the language employed does not appear to be targeted toward community members but rather toward those with significant scientific training.

Of notable significance is that all CRs reported gaining knowledge, confidence, and research skills from this course. This illustrates the value of such trainings for CRs, especially those who lack formal research backgrounds, as well as the potential for such value to increase if programs were designed or adapted to be more inclusive of community members.

While CRs reported benefitting from the tailored trainings devised by university researchers, this scaffolded approach did not fully eliminate the problems CRs experienced while working on the required human subjects course. Such a finding suggests that, while “pre-packaged” remote trainings can be a valuable addition to the preparatory phases of community-engaged initiatives, researchers should not rely exclusively on such trainings but should instead integrate this approach with other strategies that are adapted to the groups with which they are working.

Community-engaged approaches to research such as CBPR are critical in advancing health equity. Barriers which discourage community members’ involvement in conducting research should be addressed in order to help reduce health disparities, minimize attrition, promote stronger academic-community partnerships, and ensure the appropriate level of knowledge among all parties engaging with human subjects. Onerous university requirements relating to community-engaged research may impede CRs’ acquisition of such knowledge or discourage researchers from utilizing community-engaged approaches altogether.

The six CRs in the present study were all juggling familial, work, and additional non-research-related responsibilities. Although all six completed the required modules and participated fully in conducting the subsequent research project, this may not be viable for CRs in all studies and may restrict community-academic partnerships to include only select community members (e.g., those with more flexible work schedules, greater childcare options, and higher literacy levels). Such constraints may undermine one of the key motivations for and potential benefits of conducting community-engaged research.

There are a number of alternative programs that have been developed to be more responsive to community partners engaged in research. These include the University of Illinois – Chicago’s CIRTification, NYU’s Cert Program, the University of Pittsburgh’s CPRET and Certification program, and other courses which are tailored to specific groups (e.g., a training catered toward Cambodian and Lao communities).^11,24–26^ Additional research exploring community members’ experiences with these trainings can help ensure that programs provide the information needed to protect human subjects while also facilitating community members’ involvement in conducting research. The responsibility rests in part on universities to adopt, adapt, and permit the use of such courses as alternatives to current human subjects protection requirements.

While many community members are capable of successfully completing the existing requirements, others may be discouraged or deterred from participating in community-engaged research upon interacting with a system that presumes a level of scientific, semantic, and test-taking expertise which is neither universal nor needed by most community members. It is likely that the individuals most affected by this will disproportionately belong to groups which have been historically excluded from research processes.

The equity implications of existing human subjects requirements are of high importance. CBPR and other community-engaged approaches to research can help assure that often marginalized voices are heard. Treating CRs’ experiences with mandatory training requirements both thoughtfully and respectfully is essential to realizing this ambition and producing a more equitable scholarship.

This study is subject to several limitations. First, the sample size of community researchers was small (n=6), and it is possible that a larger sample would have generated different findings. Second, the individuals interviewed in this study are not representative of all community members who participate in community-engaged research and thus the findings have limited generalizability. Third, due to scheduling conflicts, many of the interviews were conducted months after CRs had completed the training. This may have influenced CRs’ recollections of their experience with the human subjects course. Fourth, half of the interviews took place prior to CRs’ interacting with human subjects whereas the other half took place after CRs had begun conducting interviews. Finally, CRs in this study completed a particular training per university requirements; however, numerous iterations of such trainings exist, as do variations in specific university requirements. Future research should aim to address these limitations through studies involving multiple institutions and human subjects protection courses.

## Conclusions

Community-engaged research is an important tool for improving community health and advancing health equity. Barriers such as currently required human subjects trainings may render this type of research unduly burdensome or impractical to carry out. This runs the risk of weakening current or potential academic-community partnerships and disincentivizing both community members and academic researchers from embarking on such projects in the future.

While there is increasing recognition of the importance of meaningful community engagement in research across all project phases, structural barriers such as university-required trainings which fail to accommodate the unique roles, experiences, and skillsets of community members must be addressed before the full potential of CBPR can be realized.

The protection of human subjects is central to the ethical conduct of research; however, less attention has been afforded to assuring that mandatory trainings are properly tailored to community members who are acting as partners in research projects. It is incumbent on universities to reexamine their human subjects training requirements in order to ensure that the ‘C’ in CBPR remains a priority.

## Data Availability

All data produced in the present study are available upon reasonable request to the authors

## Acknowledgements

The authors would like to acknowledge the Greater Newark Health Care Coalition (GNHCC), an organization that partnered with the university-based research team to recruit community researchers and study participants.

## Funding Details

There is no funding to disclose for this study. The broader CBPR project from which this sub-study is derived was funded through a grant from Merck for Mothers to GNHCC. Newark was awarded the grant as part of the Safer Childbirth Cities Initiative.

## Disclosure Statement

The authors report there are no competing interests to declare.

## Notes

### Competing Interest Statement

The authors have declared no competing interest.

### Funding Statement

This study did not receive any funding

### Author Declarations

The broader CBPR study from which this sub-study is derived was approved by Rutgers University's IRB (Pro2019002981). The present sub-study was determined to be non-human subjects research as defined under 45 CFR 46.102(d).

